# Epidemic dynamics shape variant appearance and stochastic establishment: implications for vaccination

**DOI:** 10.64898/2026.07.21.26358562

**Authors:** Maria A. Gutierrez, Julia R. Gog

**Affiliations:** Odum School of Ecology, University of Georgia, USA; Department of Applied Mathematics and Theoretical Physics, University of Cambridge, UK

**Keywords:** vaccine escape strains, antigenic evolution, stochastic establishment, Kendall formula

## Abstract

In a population model for an infectious disease, we consider the early stochastic dynamics of an emergent (“mutant”) strain, appearing and spreading during an epidemic of another (“wildtype”) strain. The mutant may not reach establishment in the host population. The time at which the mutant first appears determines its probability of establishment. We calculate this establishment probability with two methods. The first method assumes a classical branching process, with a constant transmission rate. The second method reflects the changing size of the pool of susceptible hosts, due to the dynamics of the wildtype. We find that susceptible depletion can substantially impact the establishment probability. We explore the consequences of this stochastic establishment on the “escape pressure” acting on a pathogen to produce immune escape variants. We find that the overall escape pressure rate depends strongly on the appearance time of the mutant, especially if the establishment probability is itself shaped by the continued spread of the wildtype. In most scenarios, the escape pressure rate (and thus, the risk of new escape variants) peaks slightly earlier than the prevalence of the wildtype strain. Integrating the escape pressure over time, we obtain the cumulative escape pressure generated by the wildtype epidemic. The relationship between the escape pressure and the vaccination coverage depends on the cross-immunity, due to susceptible depletion. For example, with intermediate cross-immunity, the risk of immune escape may be lowest at intermediate vaccination coverages. Thus, these results raise important considerations for vaccination strategies in response to novel outbreaks.

**Graphical abstract:** 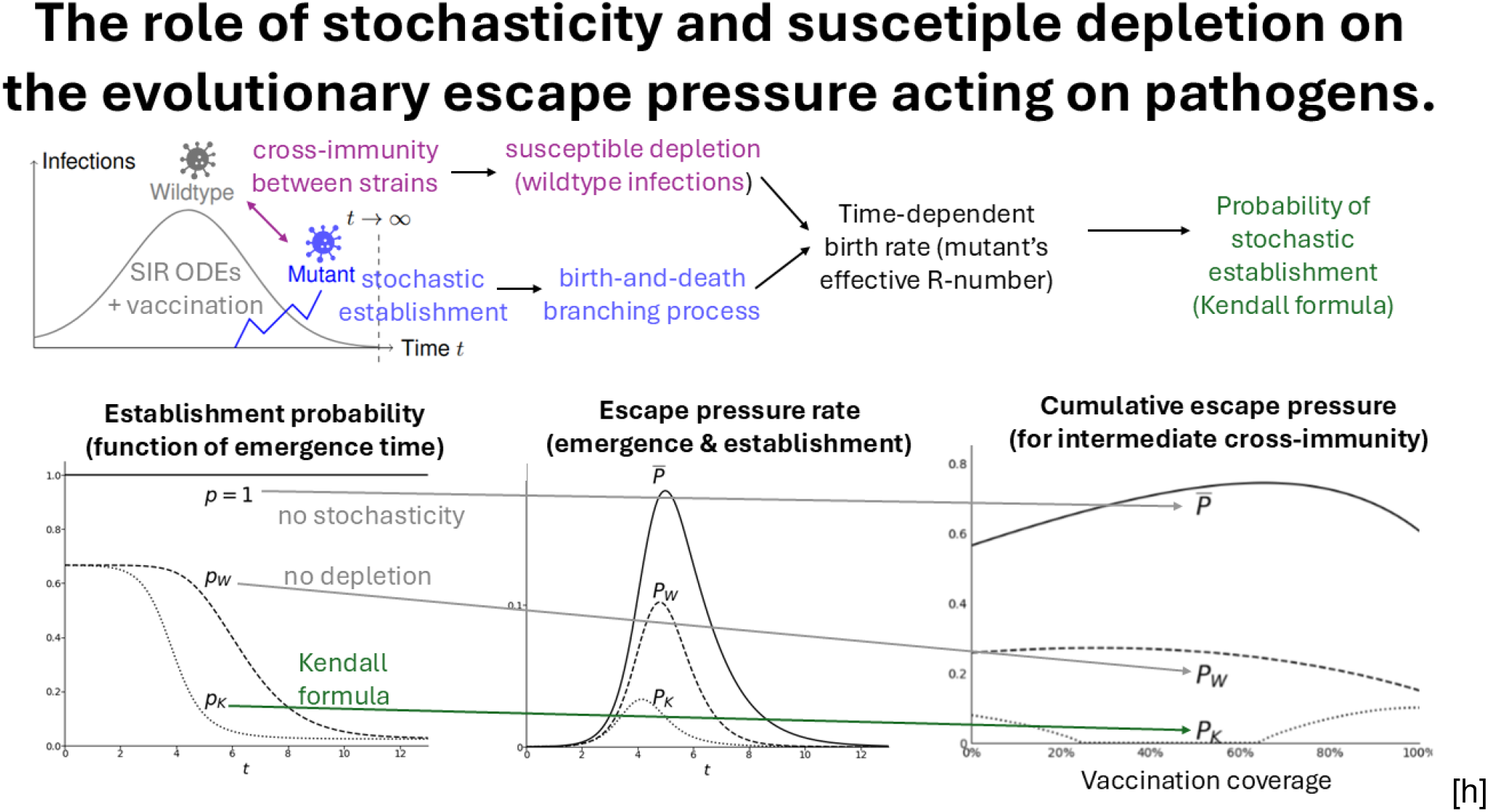

**Highlights:**

- Risk of variant emergence during epidemic wave investigated.
- Improvements on methods of understanding stochastic emergence.
- Variants appearing just before epidemic peak should be most concerning.
- Intermediate vaccine coverage may minimise risk of mutant establishment.
- Strain cross-immunity and mutant emergence timing shape these evolutionary patterns.

## 1 Introduction

The emergence of SARS-CoV-2 Variants of Concern (VoCs) that partially evade existing immunity—such as Beta [11], Delta [38], Omicron [22], and more recent (sub)variants [31]—has motivated many studies of the evolutionary consequences of vaccination [10, 14, 18, 40, 43]. At the host level, partial immunity itself can drive the natural evolution of some pathogens towards antigenic escape [17]. At the population level, the evolutionary impact of partial population immunity is even less clear [5, 17]: immunity has the potential to either reduce or increase the appearance rate of escape strains [18], and it may also facilitate their spread [6]. Here we address how the overall risk of immune escape during an epidemic wave depends on time and the vaccination coverage of the host population.

Recent works on COVID-19 vaccine escape [14, 18, 43, 47] only study the evolutionary forces driving the *appearance* of mutant (“escape”) strains. In contrast, some other works on vaccine escape [10, 24, 33, 39, 40, 52] also model the stochastic *establishment* of mutants. These two processes drive evolution at different scales and may act in conflict rather than synergy [5]. Here we explore when and how stochasticity during establishment affects the emergence—the appearance *and* subsequent establishment—of immune escape strains.

To calculate the establishment probability of a mutant strain, traditional branching theory is often used [24, 33, 39, 49]. The survival probability of a Poisson birth-death process also appears in models of immune escape that do not consider vaccination [36, 48]. Other studies consider the emergence a cross-reactive variant after a complete initial epidemic wave or while at least one other strain is endemic [36, 48]. Traditional branching theory assumes a constant pool of susceptible hosts during the phase of stochastic establishment [25]. During this initial stochastic phase, the mutant strain infects relatively few hosts, so its impact on the size of the susceptible pool is negligible [25]. So this assumption may be justified on a very short timescale or when there is no other process involved, however is unsound when there is something else changing host availability, such as such as vaccination [44] or an ongoing epidemic of the wildtype [10, 20]. Here we explore how susceptible depletion affects the establishment of an immune escape strain.

The transmission rate of the mutant is proportional to the size of its susceptible pool, so susceptible depletion will impair its establishment. Kendall’s formula [26] extends classical theory [49] by considering a branching process with *time-dependent* “birth” (transmission) and “death” (rates). Therefore, Kendall’s formula can account for changes in the susceptible pool during the stochastic establishment of the mutant strain [10]. This formula can also study establishment with time-dependent rates in other epidemiological contexts, such as during seasonal epidemics [3]. The extent to which the spread of the wildtype strain affects the pool of hosts susceptible to the mutant depends on the cross-immunity between the strains. For instance, in [10, 24, 40], hosts recovered from a wildtype infection are fully protected against the mutant strain (until immunity wanes). However, the cross-protection—from vaccination or infection—might differ widely amongst strains and vaccines. Here we consider the full range of possible values for the cross-protection to the mutant in hosts immune to the wildtype.

In this paper, we study the early stochastic dynamics of a mutant strain that appears during an epidemic of another strain (Section 2.1). We use a time-inhomogeneous birth-and-death branching process to calculate the mutant’s establishment probability while accounting for susceptible depletion (Section 2.2). We then multiply the establishment probability by the escape pressure rate of [18] to calculate the overall escape pressure for the *emergence* of escape strains (Section 2.3). We find that the emergence probability of mutants strains changes rapidly with time, as the wildtype spreads and depletes the pool of susceptibles (Section 3.1). We also find that the strain cross-immunity determines how the vaccination coverage affects the cumulative escape pressure (Section 3.2). In particular, intermediate levels of cross-immunity and vaccination lead to the lowest risk of escape.

**Table 1:** Parameters for the epidemic dynamics of the wildtype and mutant strains. The parameters of the wildtype (from θ_S_ in the top row down to 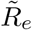) are as in [18].

| item | description |
| --- | --- |
| $\theta_S \in [0, 1]$ | relative susceptibility—to the wildtype strain—of vaccinees |
| $\theta_I \in [0, 1]$ | relative infectiousness of vaccinees (the same for both infecting strains) |
| $\theta_E \geq 0$ | relative net pathogen adaptation rate in vaccinees, as in [18] |
| $c \in [0, 1]$ | vaccination coverage of the population |
| $R_0 \geq 0$ | initial basic reproduction number of both strains (here $R_0 > 1$) |
| $R_e \geq 0$ | initial effective reproduction number of the wildtype (here $R_e > 1$) |
| $T \geq 0$ | appearance time of the mutant strain |
| $\tilde{R}_e \geq 0$ | effective reproduction number of the mutant, given by (2) |
| $\sigma \in [0, 1]$ | strain cross-immunity (protection against infection with the mutant) |
| $\eta \in [0, 1]$ | $1 - \sigma$, evasiveness of the mutant (escape from immunity to the wildtype) |

## 2 Model

### 2.1 Epidemic model

We assume that a wildtype strain generates a single epidemic wave, equivalent to the transient scenario of [18]. As in [18], *c* is the vaccination coverage and *θ*_*S*_ is the (all-or-none [19]) relative susceptibility of vaccinees (*θ*_*S*_ = 1 means no protection against infection). At any time *t*, a proportion *V* (*t*) = *c*(1− *θ*_*S*_) of the population has full protection against infection with the wildtype. We assume that the epidemic dynamics of the wildtype do not change with the possible appearance of mutant strains (we discuss this in Section 4). Hence, the SIR dynamics of [18] determine the population proportions of unvaccinated and vaccinated hosts who are susceptible (*S*_*U*_, *S*_*V*_ ); infected (*I*_*U*_, *I*_*V*_ ); and recovered (*R*_*U*_, *R*_*V*_ ). These epidemic dynamics obey

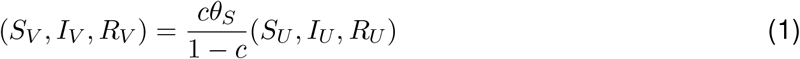

at all times [18]. The initial effective reproduction number of the wildtype is *R*_*e*_ = *R*_0_(1 − *c*(1− *θ*_*S*_*θ*_*I*_)), where *R*_0_ > 1 is the basic reproduction number of the disease. We assume *R*_*e*_ > 1 so that vaccination alone does not prevent the wildtype epidemic.

During the epidemic of the wildtype, a mutant strain may arise at any time. We assume this mutant may partially escape immunity to the wildtype strain. We use σ ∈ [0, 1] to denote the cross-immunity between the strains (if σ = 1, there is full cross-immunity), which we assume provides all-or-none protection against becoming infected. It is also convenient to define the *evasiveness* of the mutant, *η* = 1 − σ: the probability that hosts immune to the wildtype are susceptible to the mutant.

We assume that vaccination (against the wildtype) also provides the same partial protection against infection by the mutant as it does for the wildtype strain. Vaccination also reduces infectiousness by a factor *θ*_*I*_ in both mutant and wildtype infections. In addition, prior wildtype infection reduces infectiousness of the mutant by the same multiplicative factor *θ*_*I*_, so if a host has previously been vaccinated *and* had the wildtype, infectiousness is scaled by 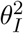 Section 4.2 discusses these assumptions.

Furthermore, we also assume that an active infection with the wildtype provides no additional protection during that infection against the escape strain. Hence, coinfections are possible: Appendix A explores an alternative scenario without coinfections.

Finally, we assume the same intrinsic transmissibility and recovery period for both strains, so that *R*_0_ is also the *basic* reproduction number of the mutant. Therefore, the *effective* reproduction number of the mutant at time *t* is

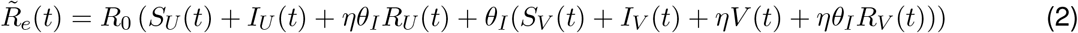

where *S*_*U*_, *S*_*V*_, *I*_*U*_, *I*_*V*_, *R*_*U*_, *R*_*V*_ and *V* obey the SIR dynamics of [18] for the wildtype epidemic. Using *V* (*t*) = *c*(1 − *θ*_*S*_), the proportionality relation (1) between the vaccinated and unvaccinated compartments, and *S*_*U*_ + *I*_*U*_ = 1 − *c* − *R*_*U*_, we obtain

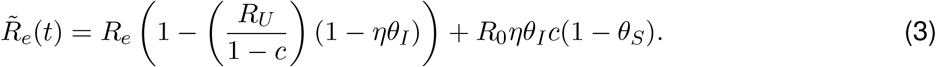

For the establishment of the mutant (below), 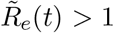 is necessary (but not sufficient). The number of (unvaccinated) recovered individuals *R*_*U*_ (*t*) increases with time, so 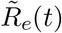 decreases monotonically throughout the epidemic (unlike in Appendix A). Since 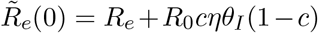 the effective reproduction number of the mutant is always greater than one (because we assume *R*_*e*_ > 1) if it appears at *t* = 0 (the start of the wildtype epidemic). However, 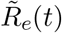 may later drop below one, preventing the establishment of the mutant. For example, at the end of the wildtype outbreak, the effective reproduction number of the mutant is 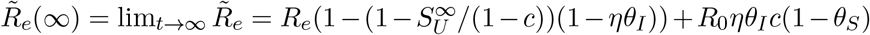 As in [18], 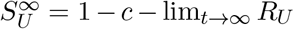 are the remaining unvaccinated susceptibles. Hence, with full cross-immunity (*η* = 0), 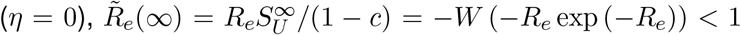 where *W* is the Lambert W function [18, 30]. In other words, a mutant strain which does not escape immunity (full cross-immunity σ = 1) and appears after the wildtype epidemic (*T*→ ∞ ) always fails at establishment, because its effective reproduction number is below one.

### 2.2 Establishment probability

Even if its effective reproduction number is greater than one 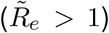, the mutant may fail to become established—due to stochastic effects when it infects only a few individuals. The probability of establishment depends on the time of appearance of the mutant, *T* .

We calculate the establishment probability using traditional branching-process theory. Assuming a single initial infection with the mutant and standard assumptions for the transmission process [25, 49], we obtain the *Whittle* probability for establishment of a mutant that appears at time *T* is

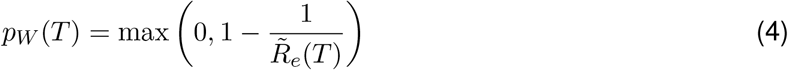

where 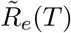is the effective reproduction number of the mutant (3). The Whittle probability (4) assumes that the birth rate of the underlying branching process (here the transmission rate, i.e., 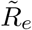) is constant during the establishment phase [25]. During this period, the mutant strain is rare in the population. Therefore, it is acceptable to ignore changes in 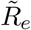 due to the spread of the mutant. In other words, the Whittle probability *pW* (*T* ) only “sees” the epidemic dynamics at the appearance time *T* . However, due to wildtype infections, the pool of individuals susceptible to the mutant might decrease quickly. Therefore,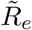 may change substantially during the phase of stochastic establishment. Hence, we now relax the assumption that the branching process has a constant birth rate (here 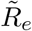 ). The death rate is still the constant recovery rate, equal to one with the chosen time units. We adapt the birth-and-death branching process with time-dependent rates of Kendall [26]. The *Kendall* establishment probability *p*_*K*_(*T* ) obeys

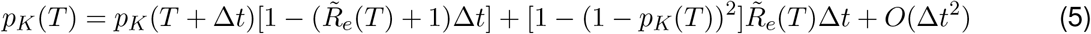

where ∆*t* is a small time interval. In the limit ∆*t* → 0, we obtain

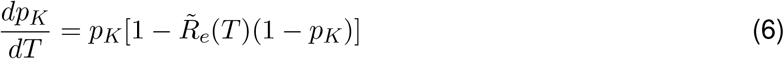

for the probability *p*_*K*_, which is always less than one (as per (9) below). A similar ODE for the establishment probability appears in [3] and [44]. Since 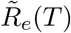 changes in time, (6) is a non-autonomous dynamical system. However, (3) gives us the trajectory of 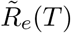 The solution for *p*_*K*_(*T* ) is (9), a version of Kendall’s formula [3, 10, 26]. To solve (6), we use a *u* = 1*/p*_*K*_ substitution and a standard integrating factor (in the ODE for *u*), obtaining

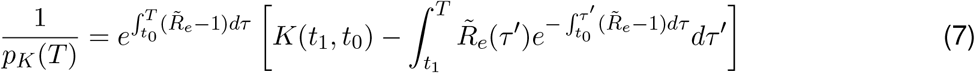

where *K*(*t*_0_, *t*_1_) is an integration constant which depends on the arbitrary times *t*_0_, *t*_1_ ≥ *T* . We set *t*_0_, *t*_1_ = ∞ and use integration by parts to get

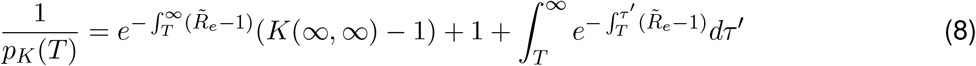

for a constant *K*(∞,∞ ). If 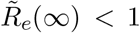 the right-most term of (8) diverges, so *p*_*K*_(*T* ) = 0 for all possible appearance times. This means that—with the Kendall formulation (6)—the mutant never reaches establishment if it cannot spread after the wildtype epidemic 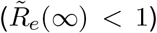 For example, *p*_*K*_ = 0 if there is full cross-immunity between both strains. If 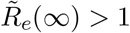 and *T* < ∞, 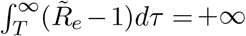 Thus, the first term of (8) vanishes and the Kendall establishment probability is

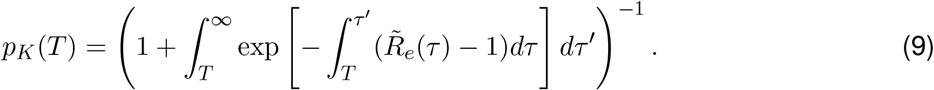

This expression shows that the values 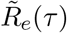 at times *τ* > *T* (i.e., after the mutant appears) influence the establishment probability *p*_*K*_(*T* ). If 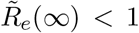 the outer integral of (9) diverges, so we recover *p*_*K*_(*T* ) = 0, as already derived above from (8). For constant 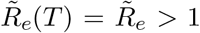 it is straight-forward to integrate (analytically) the Kendall probability (9) and recover the Whittle probability (4): 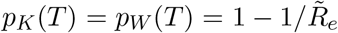 In other words, when 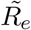 is constant, the Whittle and Kendall establishment probabilities are equal, because both arise from the same time-homogeneous birth-and-death branching process. For general 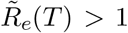 the Kendall probability (9) also approaches the Whittle probability in the limits of *T*→ 0 (the mutant appears before the wildtype epidemic) and *T*→ ∞ (the mutant appears after the wildtype epidemic).

#### Limits of the Kendall probability

To prove that 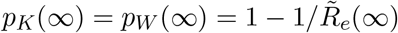 for 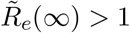 we first write

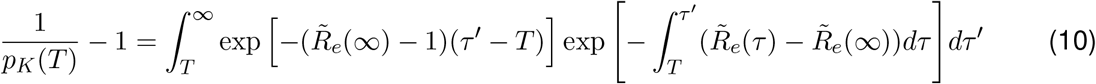

Since 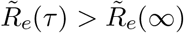 for all τ < ∞ (from Section 2.1, 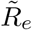 decreases monotonically),

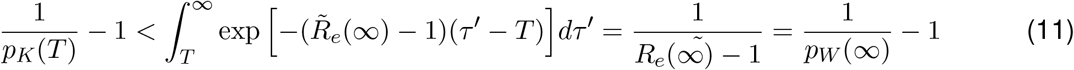

for any *T* < ∞. Moreover,

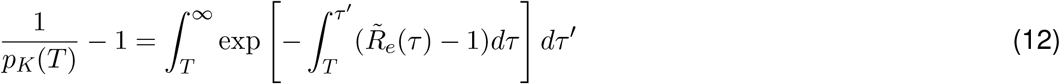

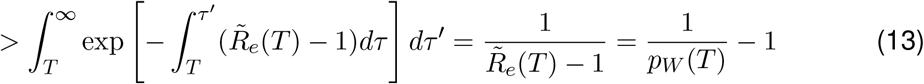

Together (11) and (13) show that *p*_*W*_ (∞) < *p*_*K*_(*T* ) < *p*_*W*_ (*T* ) for any finite appearance time (*T* < ∞). Taking *T*→∞ shows that *p*_*K*_( ) = *p*_*W*_ (∞ ). This proof assumes that 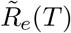 is monotonically decreasing (as shown in Section 2.1). We now consider briefly an alternative scenario which appears in Appendix A. Suppose that there exists *T*_1_ such that for all *T* > *T*_1_ sufficiently large, 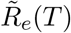 *increases* up to 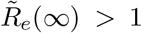 Inequalities similar to (11) and (13) show that *p*_*W*_ (*T* ) < *p*_*K*_(*T* ) < *p*_*W*_ (∞) for all *T* > *T*_1_. In particular, *p*_*K*_(∞) = *p*_*W*_ (∞) again. A more formal analysis (starting from (10)) is required to prove *p*_*K*_(∞) = *p*_*W*_ (∞) if instead 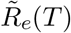converges to 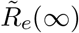 in an oscillatory manner 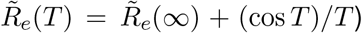 Since this scenario is far-fetched (in the context of epidemic modelling), we do not consider it further.

It is intuitive that *p*_*K*_(∞) = *p*_*W*_ (∞ ). At *T* → ∞ the wildtype strain is no longer in the host population, so it does not cause susceptible depletion. Hence, the Kendall and Whittle probabilities are equal. For instance, if 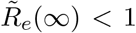 the mutant has no chance at establishment: *p*_*K*_(∞) = 0 = *p*_*W*_ (∞ ), as already shown directly from (8).

The Kendall and Whittle probabilities also approach the same value *p*_*K*_(0) = *p*_*W*_ (0) at early times (*T*≈ 0) if 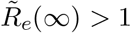. The mutant’s effective reproduction number 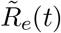 barely decreases during the very early phases of the epidemic. Hence, the mutant has enough time to reach establishment before the depletion of susceptible hosts becomes substantial. Lowering the fraction (*ϵ ≪* 1) of hosts that initiate the wildtype epidemic gives the mutant more time to reach establishment without susceptible depletion, recovering the time-homogeneous branching process that leads to the Whittle probability (4). It can be shown from (9) that if 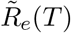 is constant at all times *T* < *T*_0_, then 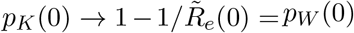 as *T*_0_ → ∞, provided that 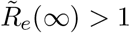

### 2.3 Escape pressure

We use an escape pressure function [14, 18] to represent the combined risk of both appearance *and* stochastic establishment of escape strains (at the population level). We assume that the appearance rate of these strains is proportional to the escape pressure rate of [18]: *P* (*t*) = *I*_*U*_ (*t*) + *θ*_*E*_*I*_*V*_ (*t*). We refer to *θ*_*E*_ as the relative net pathogen adaptation rate [17] in vaccinees. The parameter *θ*_*E*_ may take any value from zero (only infections in the unvaccinated contribute to the escape pressure rate) to infinity (only infections in the vaccinated contribute to the escape pressure rate) [18]. In this limit *θ*_*E*_→ ∞, the escape pressure rate *P(t)* is unbounded. However, we are primarily interested in the relative value of *P* (*t*) [18], so here we use the normalisation

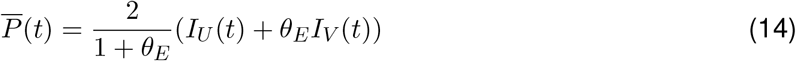

The factor of 2 in (14) means that if *θ*_*E*_ = 1 (so the net pathogen adaptation rate is the same for all hosts), the escape pressure rate is equal to the total prevalence: 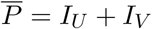 .

The *overall* evolutionary potential for the appearance and establishment of escape strains is the product of the escape pressure rate 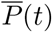 and *p*_⊗_ (⊗ = *K, W* ), one of the establishment probabilities. We use = *W* for the (traditional) Whittle process (4) and ⊗ = *K* for the (time-inhomogeneous) Kendall process (9). Each formulation leads to a different overall escape pressure *rate*:

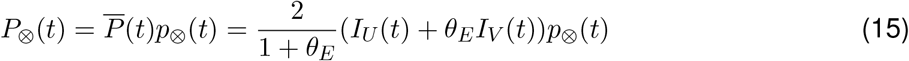

For ⊗ = *W, K*. Expression (15) measures the pathogen adaptation rate at the population level: in the Whittle formulation (⊗ = *W* ), *P*_*W*_ (*t*) is similar to the “phylodynamic” net adaptation rate that appears in [36] (which only considers an endemic equilibrium and no vaccination). The escape pressure rate (15) is also similar to the time-dependent rates in [10] (which uses ⊗ = *K*) and [24] (which uses ⊗ = *W* ), but (15) does not give a true probability for the establishment of a vaccine escape mutant (see the discussion in Section 4). Section 3.1 studies how *p*_⊗_(*t*) and *P*_⊗_(*t*) change over time.

We also consider the cumulative escape pressure: the integral of (15) over the wildtype epidemic wave,

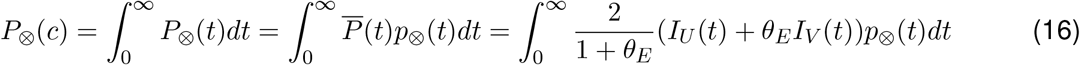

for ⊗ = *W, K*. As in [18], the escape pressure (16) depends on the vaccination coverage *c*. We integrate (16) numerically. In Section 3.2, we compare the escape pressures *P*_⊗_(*c*) with

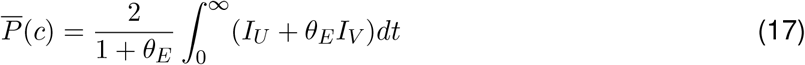

i.e., the same integral (16) as *P*_⊗_(*c*), except with *p*_⊗_ = 1 at all times. Except for the multiplicative normalisation, 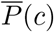 is the cumulative escape pressure *P*_*a*_(*c*) of [18]. Unlike in [18], here we do not normalise the cumulative escape pressures 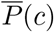 and *P*_⊗_(*c*) by their value in the absence of vaccination (*c* = 0), because *P*_*K*_(0) may be zero.

## 3 Results

### 3.1 Establishment probability and escape pressure rate

First, we consider the escape pressure rates *P*_⊗_(*t*) and the establishment probabilities *p*_⊗_(*t*) (Figure 1). We compare the overall rates *P*_⊗_(*t*) with the escape pressure rate 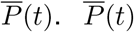 does not account for the possible failure of mutant establishment, so its maximum corresponds to the peak in prevalence of the wildtype strain [18]. We find that the establishment functions, *p*_⊗_(*t*) and *P*_⊗_(*t*), have one of two general behaviours. We explain these behaviours below. No other general behaviours are possible here—because 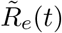 is monotonically decreasing and, thus, so are the probabilities *p*_⊗_(*t*)—but a third behaviour appears in Figure A1 (because a non-monotonic *R*_*e*_(*t*) is possible in Appendix A). Figure 1 uses representative epidemiological parameters.

**Figure 1:**
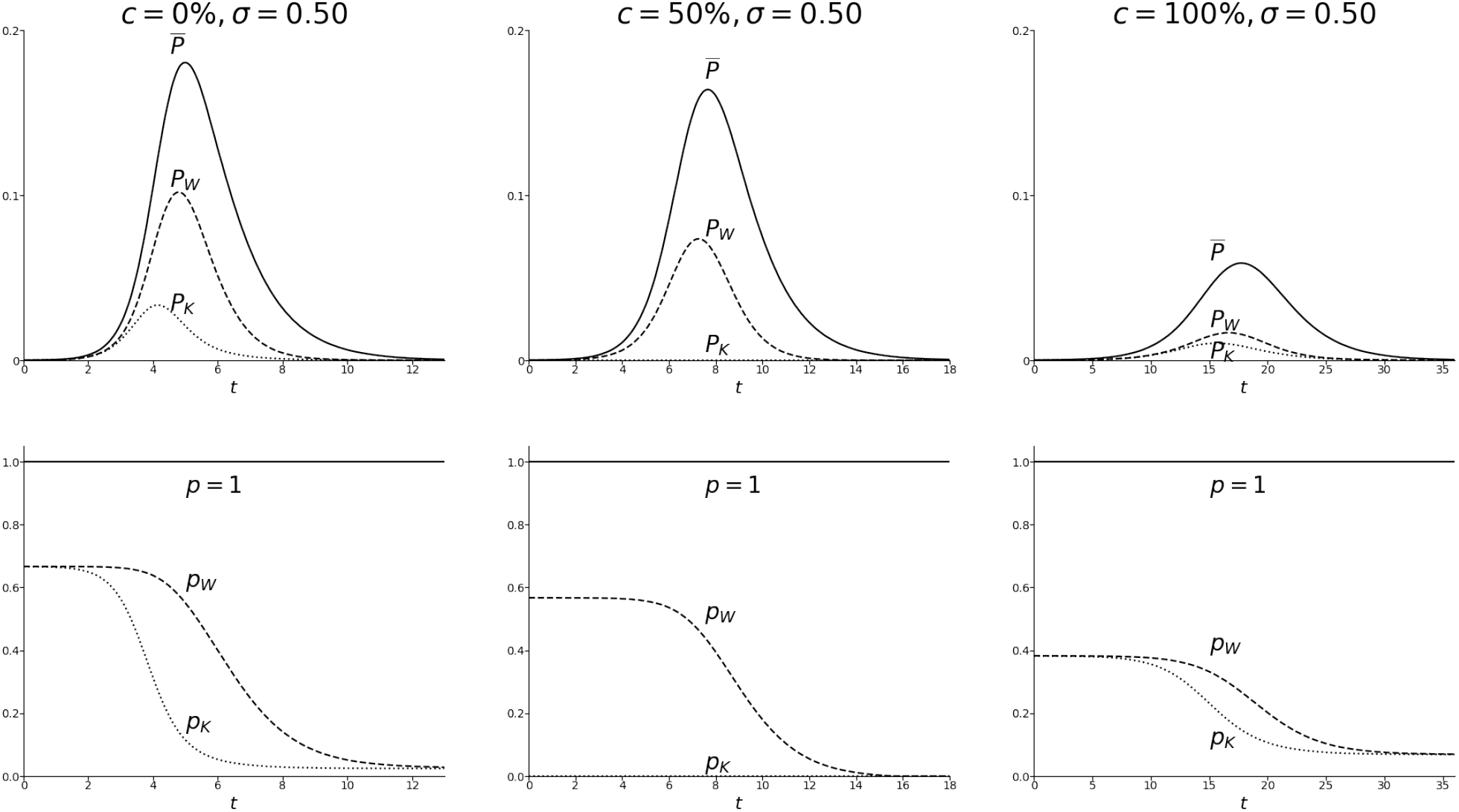
Top row: escape pressure rates P_⊗_(t) and 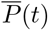. Bottom row: establishment probabilities p_⊗_(t) (and, for comparison, p = 1). The difference between the columns is the vaccination coverage c, which increases from left (no vaccination) to right (fully vaccinated population). The choice of θ_E_ does not change the qualitative behaviour of the escape pressure rates under the SIR model of [18], because I_U_ (t) = cθ_S_I_V_ (t)/(1 − c). Fixed R_0_ = 3, θ_S_ = 0.8, θ_I_ = 0.6 (representative parameters chosen to show all possible qualitative patterns across the manuscript without changing these parameters between the figures), and σ = 0.5, θ_E_ = 7/3 (corresponding to the third plot in Figure 2).

Figure 1 demonstrates that there is a narrow time window during which mutants are likely to both appear and succeed at (stochastic) establishment. On the one hand, at early times, mutants are unlikely to appear; but if they do, they are relatively likely to reach establishment. On the other hand, at later times, mutants may not be able to spread. Figure 1 also shows how *changes* in the susceptible pool due to the wildtype may substantially impact the mutant: for some parameters, establishment is impossible, regardless of the appearance time (*p*_*K*_ = 0, versus *p*_*W*_ > 0 without susceptible depletion).

In the left and right columns of Figure 1, the establishment probabilities *p*_⊗_ monotonically decrease with time. Even after the wildtype epidemic is over, enough susceptible hosts remain, allowing the mutant to spread 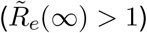 Therefore, the mutant always has a chance at establishment (*p*_⊗_(*t*) > 0). However, the possibility of unsuccessful establishment substantially reduces the escape rates *P*_⊗_(*t*) relative to 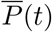, their deterministic counterpart. Moreover, the *P*_⊗_(*t*)s peak slightly earlier than 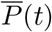 because the establishment probabilities *p*_⊗_(*t*) decrease throughout the epidemic. The Kendall probability *p*_*K*_(*t*) decreases earlier than the Whittle probability *p*_*W*_ (*t*), because the expression (9) for *p*_*K*_ “foresees” the upcoming decrease in 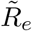 Therefore, the peak of the Kendall escape pressure rate *P*_*K*_(*t*) is lower and appears earlier in the epidemic, relative to *P*_*W*_ (*t*). As proven with (13), the Kendall probability is lower than the Whittle probability (*p*_*K*_ < *p*_*W*_ ) at all finite positive times. (Appendix A discusses alternative assumptions for the mutant, such that *R*_*e*_(*t*) is not always decreasing, and, thus, *p*_*K*_(*t*) > *p*_*W*_ (*t*) is possible.) Both probabilities *p*_*K*_(*t*) and *p*_*K*_(*t*) approach the same value once the wildtype is no longer present (*p*_*K*_(∞) = *p*_*W*_ (∞) as proven in Section 2.2). Since there is no additional susceptible depletion at late times (*T*→ ∞ ), 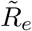 is constant and the Kendall formulation recovers the classical time-homogeneous branching process (which leads to the Whittle probability).

The middle column in Figure 1 exemplifies a more extreme consequence of *p*_*K*_(*T* )’s ability to foresee 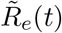 at future times *t* > *T* (after appearance of the mutant). The effective reproduction number of the mutant, 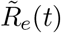 decreases with time, dropping below one before the epidemic ends. From this time onwards, since 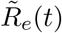 is decreasing, 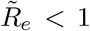 Thus, the Whittle probability *p*_*W*_ (*t*) and the corresponding escape pressure rate *P*_*W*_ (*t*) are zero at late times. Before reaching the threshold 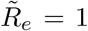 *p*_*W*_ behaves as in the other columns in Figure 1 (i.e., it decreases with time). Hence, *P*_*W*_ is also as discussed above. However, the Kendall escape pressure rate vanishes at all appearance times (*P*_*K*_(*t*) = 0), because the Kendall establishment probability is always zero (*p*_*K*_(*t*) = 0). With the Kendall formulation, if the mutant cannot spread at late times (that is, 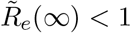 ) establishment is impossible (regardless of the appearance time). Section 4 discusses this result.

At *T* = 0, there is a notable—but ultimately irrelevant—difference between the two aforementioned scenarios. As explained above, if 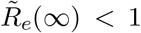 the Kendall probability is zero even at the start of the wildtype epidemic (*p*_*K*_(0) = 0). However, if 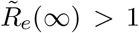 the Kendall and Whittle approach the same value *p*_*K*_(0) ≈ *p*_*K*_(0) > 0 for *T* ≈ 0 (as explained at the end of Section 2.2). However, these very early values of the establishment probabilities have no meaningful consequences (at least in the context of this chapter). Since there are essentially no infections at early times, mutants cannot appear 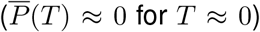 Hence, the Whittle and Kendall escape pressure rates (15) are both zero (*P*_⊗_(*t*) = *P* (*T* )*p*_⊗_(*T* ) ≈ 0), no matter the value of the establishment probabilities *p*_⊗_(0).

For 0 < *T* <∞, the Kendall escape pressure rate is lower than or equal to the Whittle escape pressure rate (*P*_*K*_(*T* ) ≤ *P*_*W*_ (*T* )), with equality if and only if 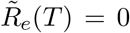 This result follows directly from *p*_*K*_(*T* ) ≤ *p*_*W*_ (*T* ), which we proved with (13) under the assumption that 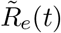 monotonically decreases (see Appendix A for a counterexample).

### 3.2 Overall cumulative escape pressure

We now focus on how the vaccination coverage *c* affects the cumulative escape pressures *P*_⊗_(*c*) and 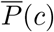 (Figures 2 and 3). We choose parameter values across the cross-immunity range σ∈ [0, 1] to display all possible qualitative patterns. The relative net pathogen adaptation rate in vaccinees, *θ*_*E*_, determines the shape of 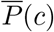 as a function of the vaccination coverage [18]. Figures 2 and 3 are representative examples of the two possible behaviours for 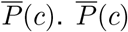 is unimodal in Figure 2 (with *θ*_*E*_ = 7*/*3) and monotonically decreasing in Figure 3 (with *θ*_*E*_ = 1).

**Figure 2:**
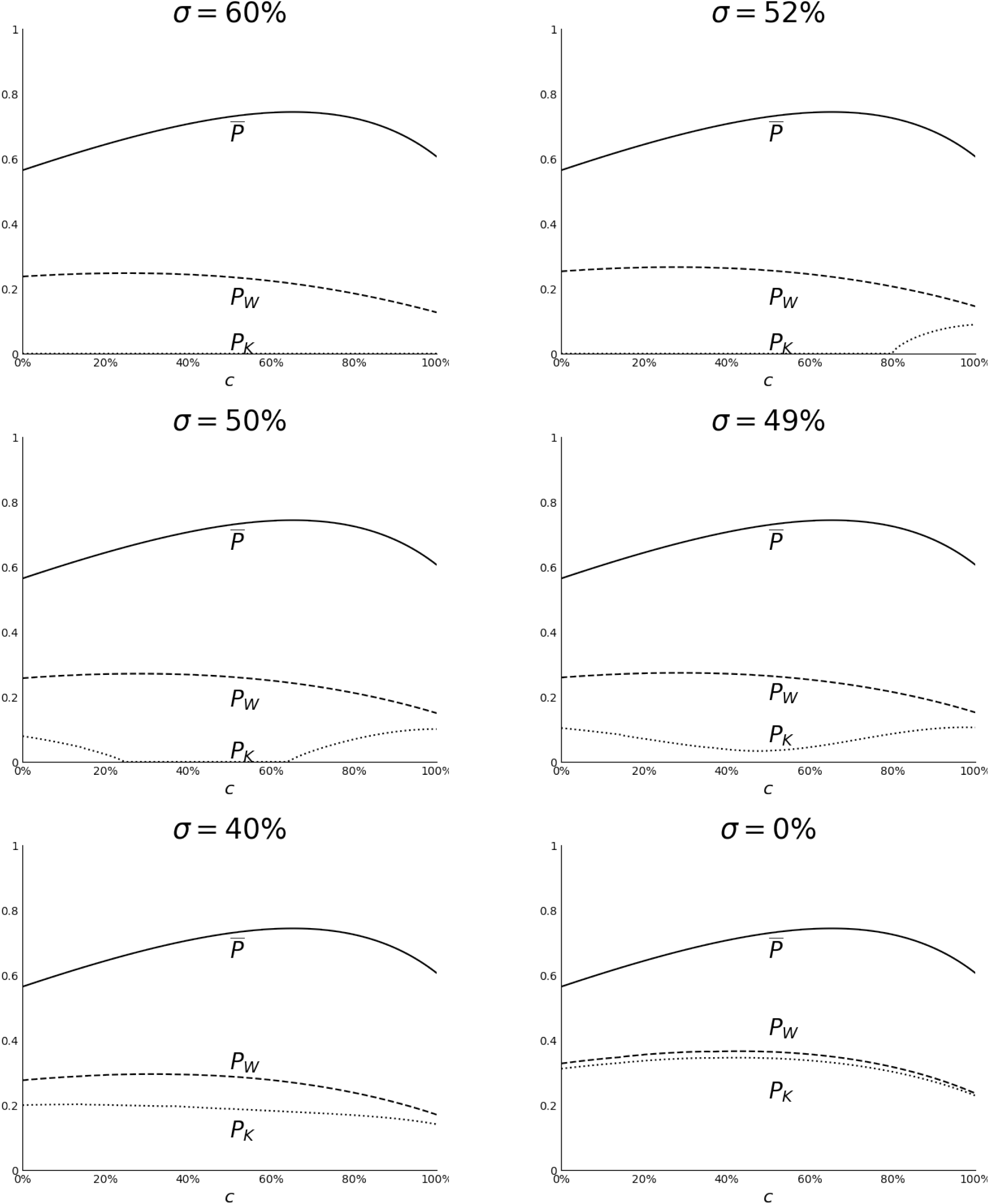
Cumulative escape pressures P_⊗_(c) and 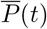 for different values of the cross-immunity σ. Here θ_E_ = 7/3 so that 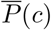is unimodal. The cross-immunity σ, decreases (increasing immune evasiveness η = 1 − σ) from the top left corner to the bottom right corner. Plots with cross-immunity σ > 60% (not shown) are qualitatively the same as the plot with σ = 60% (top right). Fixed R_0_ = 3, θ_S_ = 0.8, θ_I_ = 0.6 (as in Figure 1) and θ_E_ = 7/3.

**Figure 3:**
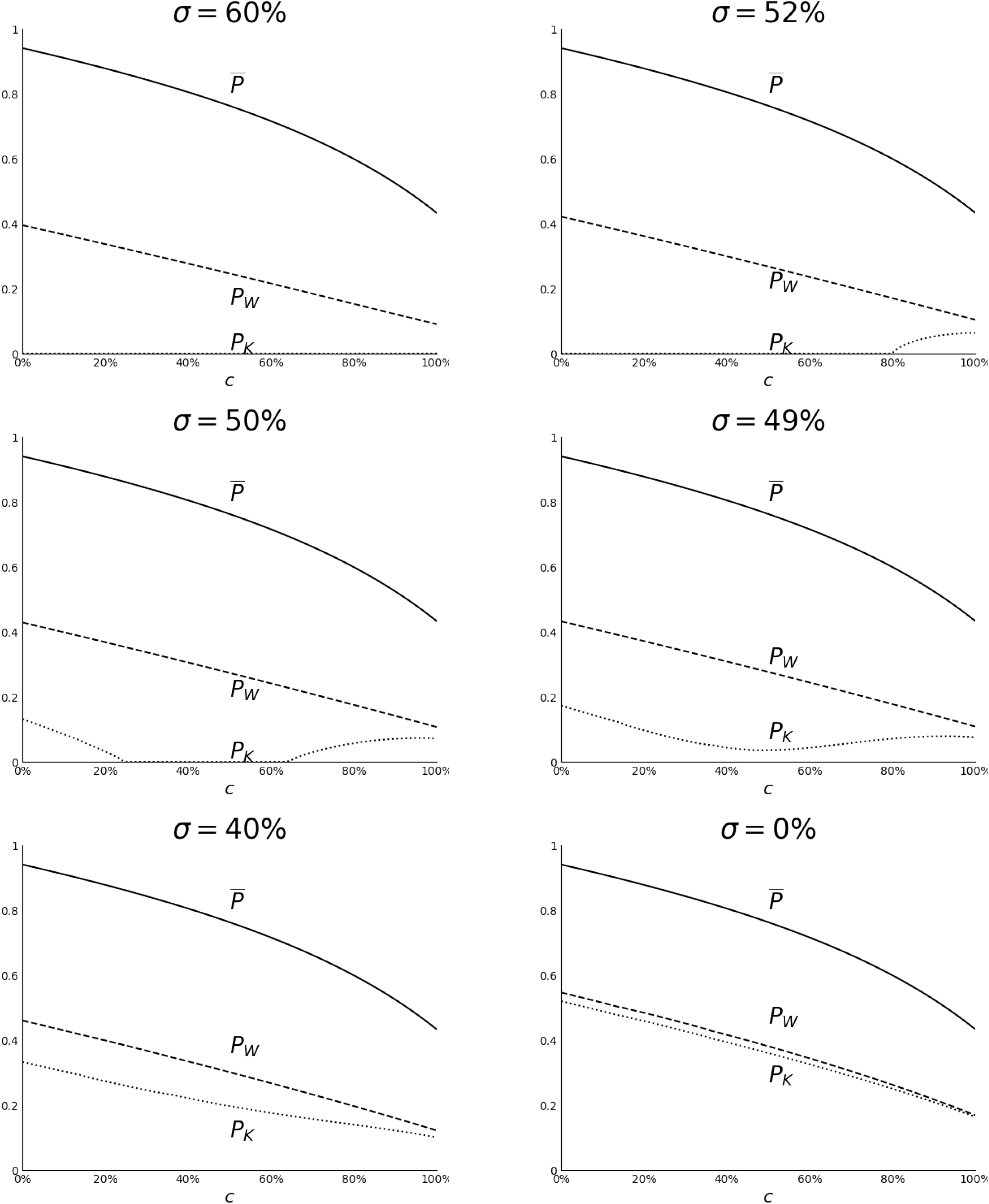
Cumulative escape pressures P_⊗_(c) and P (c) as in Figure 2 but with θ_E_ = 1, so that 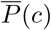 is decreasing. The cross-immunity σ, decreases (increasing immune evasiveness η = 1 − σ) from the top left corner to the bottom right corner. Plots with cross-immunity σ > 60% (not shown) are qualitatively the same as the plot with σ = 60% (top right). FixedR_0_ = 3, θ_S_ = 0.8, θ_I_ = 0.6 (as in Figures 1 and 2) and θ_E_ = 1.

Using the escape pressure 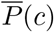of [18] as a baseline, we infer from Figures 2 and 3 general trends about *P*_*W*_ (*c*) and *P*_*K*_(*c*). On the one hand, the Whittle establishment probability (4) generally does not change substantially the shape of the Whittle escape pressure *P*_*W*_ (*c*), relative to the baseline 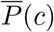 On the other hand, the Kendall establishment probability (9) may lead to different patterns in the Kendall escape pressure *P*_*K*_(*c*). The strain cross-immunity determines how *P*_*K*_(*c*) depends on the vaccination coverage (see Table 2 and next paragraph). Regardless of the cross-immunity, *P*_*K*_(*c*) < *P*_*W*_ (*c*), because *P*_*K*_(*t*) ≤ *P*_*W*_ (*t*) (with equality only at some times, as per the last paragraph of Section 3.1).

**Table 2.** The cross-immunity to the mutant strain, σ = 1 − η, determines how the cumulative escape pressure P_K_(c) (which uses a time-inhomogeneous branching process for establishment) depends on the vaccination coverage c. In contrast, the cross-immunity is unimportant for the qualitative patterns of the escape pressure P_W_ (c) (which uses a traditional branching process). P_W_ (c) has the same shape as 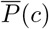 the escape pressure from [18] (which does not consider strain establishment).

| cross-immunity $\sigma$ | pattern of the escape pressure $P_K(c)$ |
| --- | --- |
| full (no escape) | zero at all vaccination coverages $c$ |
| high | increasing at high vaccination coverages, zero otherwise |
| intermediate | U-shape (lowest—possibly zero—at intermediate coverages) |
| low or none | as $\bar{P}(c)$ and $P_W(c)$: unimodal (high $\theta_E$) or decreasing (otherwise) |

If the cross-immunity σ is low (bottom rows in Figures 2 and 3), the escape pressure *P*_*K*_(*c*) has the same qualitative shape as 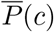 and *P*_*W*_ (*c*) (i.e. unimodal in Figure 2 and decreasing in Figure 3). However, *P*_*K*_(*c*) is qualitatively different for intermediate or high cross-immunity σ (i.e. low escape *η*). With full cross-immunity (σ = 1), the mutant always fails at establishment (as in the middle column in Figure 1). If there is some immune evasion (low *η* > 0, i.e, high σ < 1), the mutant may only reach establishment at high vaccination coverages (top right in Figures 2 and 3), because it needs these vaccinated hosts to spread (as in the right column in Figure 1). With slightly more immune evasion (lower cross-immunity), the mutant may spread at low or high vaccination, but not for intermediate levels (Figures 2 and 3: middle row, left). If the vaccination coverage is low, the wildtype infects many unvaccinated hosts, and the mutant can later spread through them (left column in Figure 1). Therefore, the overall escape pressure decreases for low coverages, reaches zero at intermediate vaccination levels, and later increases for high vaccination levels. If we reduce further the cross-immunity, intermediate vaccination levels allow establishment, but the escape pressure is lowest at these intermediate levels (Figures 2 and 3: middle row, right). In other words, the escape pressure decreases (as the vaccination coverage increases from zero) until reaching its (global) minimum at intermediate vaccination, and afterwards increases for higher vaccination levels.

## 4 Discussion

### 4.1 Overview of key results

This work shows how the spread of the wildtype strain hinders the establishment probability of mutant strains. The overall escape pressure rate changes substantially over time (Figure 1). Integrating the overall escape pressure rate over time, we obtain the cumulative escape pressure across the full epidemic period. The difference from [14, 18, 43] is that the escape pressure here measures whether mutant escape strains that appear in the population have the potential to reach establishment (and the probability thereof). Compared to [10, 24, 39], a key finding here is that the cross-protection to mutant strains from immunity to the wildtype strongly affects the cumulative escape pressure (Table 2). Surprisingly, with intermediate cross-immunity levels, the escape pressure is *lowest* at intermediate vaccination coverages. This minimum at intermediate vaccination coverages is reminiscent of the final-size results of [50] for a seasonal influenza outbreak that includes a vaccine escape strain.

Generally, early in the epidemic, mutant strains are unlikely to appear (because there are few wildtype infections that might result in mutations). Moreover, late in the epidemic few hosts remain susceptible (to infection with the mutant). Therefore, we find that the escape pressure rate peaks slightly earlier than the total prevalence of wildtype infections. We calculate the establishment probability of the mutant using a time-inhomogeneous birth-and-death branching process [26]. The Kendall establishment probability (9) reflects changes in the pool of susceptible hosts due to the spread of the wildtype *after* appearance of the mutant [10]. Thus, the resulting escape pressure *P*_*K*_(*t*) for the appearance and establishment of escape strains is lower than *P*_*W*_ (*t*): its counterpart resulting from traditional branching theory approaches (4).

Regardless of the assumptions for the establishment process, here the cumulative escape pressure (integrated over time) is lower than in [18], because mutants may fail to reach establishment. The Whittle approach for establishment (4) does not change qualitatively how the vaccination coverage affects the escape pressure, relative to the results of [18]. In other words, the relative pathogen adaptation rate in vaccinees determines the shape of *P*_*W*_ (*c*): unimodal or decreasing. The same patterns appear in the escape pressure *P*_*K*_(*c*), which results from the Kendall probability (9), *if* cross-immunity is relatively low (under 45% for the parameters we use). However, stronger cross-immunity leads to new shapes in the Kendall escape pressure *P*_*K*_(*c*).

If cross-immunity is complete, the escape pressure *P*_*K*_(*c*) is always zero (because mutants do not “escape” any immunity). If cross-immunity is partial but high, the escape pressure is zero for most vaccination coverages *c* and increasing with high vaccination coverages. For intermediate cross-immunity levels, the escape pressure *P*_*K*_(*c*) is lowest at intermediate vaccination coverages. High and low vaccination coverages result in more hosts who have partial immunity: vaccinees and recovered hosts, respectively. Instead, with intermediate vaccination levels, there are not enough partially immune hosts to facilitate the spread of the mutant. The difference in the amount of partially immune hosts (due to vaccination and/or wildtype infection) is key *if* there are few of these hosts, i.e., with intermediate cross-immunity. With intermediate cross-immunity, the establishment probability dominates the escape pressure (antigenic escape is “selection-limited” [53]). With lower cross-immunity the mutant is able to spread more easily, so the rate of appearance of escape strains dominates the overall escape pressure (which is “mutation-limited” [53]).

### 4.2 Cross-immunity assumptions

To reduce the number of model parameters, we make several assumptions (which are not necessary for the general modelling approach of this work) about how different types of immunity protect against the mutant strain.

First, we assume that immunity against wildtype infections partially protects hosts against mutant infection, and the strength of this protection—the cross-immunity σ—does not depend on the source of immunity (vaccination or wildtype infection). These assumptions follow [1] and [45], aiming to represent strain-specific (humoral) immunity in Influenza A [8] and SARS-CoV-2 [38]. We model this type of immunity as all-or-none (although leaky immunity would not change the initial effective reproduction number of the mutant strain), so that hosts immune to the wildtype are also immune to the mutant with probability σ. As noted above, we assume that this probability is the same for both vaccine- and infection-induced wildtype immunity. However, this assumption does *not* mean that vaccines protect hosts against wildtype and mutant infections with the same probability. A vaccinee is immune to the mutant with probability σ *if* they are also immune to the wildtype. Since vaccination does not elicit wildtype immunity in all vaccinees (only in a proportion *θ*_*S*_, as in [18]), hosts are less likely to become immune to the mutant after vaccination (the net probability is (1 − *θ*_*S*_)σ) than after wildtype infection (probability σ, since we assume full protection against wildtype reinfection). The premise for this lower vaccine efficacy against the mutant is that vaccine-induced antibodies do not neutralise any additional epitopes in the mutant (relative to the wildtype).

Second, we model “broad” immunity—conserved across antigenic space—as follows: (i) the vaccine reduction in host infectiousness (given by 1 − *θ*_*I*_) is conserved across antigenic space (i.e., it is equally effective against both strains), (ii) infection with the wildtype reduces host infectiousness (during infection with the mutant) by the same rate as vaccination, and (iii) this infectiousness reduction is multiplicative, so that vaccinees that recover from the wildtype transmit the mutant strain at an even lower rate (due to hybrid immunity). These assumptions are motivated by SARS-CoV-2 [35, 46]. Broad immunity from (non-universal) influenza vaccines is relatively weak [21], so both [1] and [45] instead assume that only infections elicit the aforementioned reduction in host infectiousness.

Third, we ignore any possible reduction in the infectious period due by prior infection [27, 37] or vaccination [28]. This type of cross-immunity may be important for the stochastic establishment of the mutant [41] and certainly for persistence during epidemic decline [16].

Fourth, we also ignore the waning of immunity and immune imprinting, and assume no antibody-dependent enhancement (ADE) or pathogen life-history evolution. Importantly, we assume that antigenic escape does not cause costs in pathogen fitness (any such “evolutionary constraints” in the mutant may prevent its establishment [13, 29]). However, it would be easy to consider a mutant with a different basic reproduction number [10], or possibly other life-history trait values [2, 9].

Finally, we have assumed so far (as in [51]) that an active infection—with the wildtype strain—does not reduce susceptibility to the mutant. In Appendix A we instead assume that coinfections are impossible (as in [10, 24, 40]). We find that this change in assumptions makes no substantial difference in the cumulative escape pressure. In contrast to [51], we assume that during coinfections the transmission rate of the mutant strain is the same as in mutant-only infections: the wildtype strain only interferes with the mutant through the (post-infection) cross-immunity. We expect a relatively low number of coinfections (since at any time, there will not be many hosts infected with the wildtype and thus with potential for coinfection), it is perhaps not surprising that this assumption is of little consequence.

### 4.3 Limitations and implications for stochastic establishment

In addition to the assumptions for the wildtype epidemic (discussed in [18]) and the strain cross-immunity (discussed above), this work is subject to additional caveats, due to the model for establishment and the resulting escape pressure.

Unlike [10] and [24], we do not consider the long-term strain dynamics (e.g. replacement of the wildtype) following mutant establishment. This is relevant for public health because the establishment of the mutant does not guarantee that it causes a substantial public health burden. As [39] points out, the Whittle establishment probability (4) is strongly correlated with the final size of an SIR epidemic, so the Whittle probability may serve as a measure of the epidemic burden resulting from the mutant. However, this approximation may fail with the Kendall probability and ignores the wildtype-mutant interactions. In general, modelling the long-term epidemic dynamics would require considering how the mutant affects the wildtype strain. Before establishment, the mutant infects very few hosts, so we expect its impact on the epidemic dynamics of the wildtype to be negligible (thus, we do not consider it). However, *after* reaching establishment, the mutant may substantially interfere with the wildtype, requiring more complex models.

We also assume that there is no further evolution: allowing the “evolutionary rescue” of the mutant strain could lead to the establishment of a different—fitter—escape strain [29]. This work also assumes that transmission of the mutant follows a Poisson distribution (the mean of which is the effective reproduction number of the mutant). However, individual variation in transmission patterns may have a substantial effect on the establishment probability [32]. The establishment probabilities assume that the mutant initially infects a single individual, albeit this assumption is straightforward to modify [25]. Furthermore, the escape pressure here is not a real probability—unlike in [10] and [24]—for the appearance and establishment of escape strains, but rather an approximate measure of the strength of these evolutionary processes (which are not separate from each other [15]).

#### Relevance, implications, and limitations of the Kendall formulation

As introduced in Section 4.1, the Kendall formulation—based on a time-inhomogeneous birth-and-death branching process for the stochastic establishment of the mutant strain—leads to novel patterns in the escape pressure. Here we discuss its potential shortcomings.

The Kendall establishment probability *p*_*K*_(*T* ) may be zero even if the mutant’s effective reproduction number 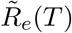 is greater than one at its appearance time *T* . In particular, *p*_*K*_(*T* ) = 0 at *all* appearance times *T* if the limit value (as *T*→ ∞ ) of the effective reproduction number of the mutant is less than one 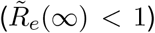 In other words, if the mutant cannot spread *after* the epidemic of the wildtype, establishment is also impossible—according to the Kendall probability—at any prior time. The time-inhomogeneous branching process (6) uses the values of 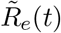 through all times *t* > *T* after mutant appearance (at time *T* ) to calculate the probability of establishment *p*_*K*_(*T* ). However, in real epidemics, mutant establishment might be fast, so that values of 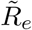 long after the mutant’s appearance should not affect the establishment probability (penalising the mutant). In other words, early establishment of the mutant should be possible, even if the wildtype eventually depletes the pool of susceptible hosts. Hence, here the Kendall probability could underestimate what one might consider the actual establishment probability. In contrast, the traditional Whittle probability is an over-estimate, because it entirely neglects the susceptible depletion from the wildtype infections that occur shortly after the mutant appears. Thus, the actual establishment probability lies between the Whittle and Kendall establishment probabilities. Depending on the wider epidemiological context, one formulation—Whittle or Kendall—may be more suitable than the other.

If the epidemiological context is such that strain establishment can be considered equivalent to reaching a large enough number of infections (for example, if there is risk of strain spillover into other populations), the Whittle probability may be more suitable at early times of the original epidemic. Before the wildtype has spread widely, the Kendall probability strongly penalises the mutant for susceptible depletion that happens much later. Therefore, a mutant that appears early might reach establishment before the wildtype grows. However, at intermediate times (when the wildtype prevalence is high), there is a quick depletion of susceptible hosts, which we need to account for, thus warranting the use of the Kendall probability. We can use these observations to justify using the Kendall escape pressure. The overall escape pressure rate (15) multiplies the establishment probability by the prevalence of the wildtype strain—technically, by a linear combination of the wildtype prevalence in unvaccinated and vaccinated hosts—which is low at the beginning of the epidemic. Therefore, both the Kendall and Whittle escape pressure rates are very close to zero at early times. These escape pressure rates only meaningfully differ when the wildtype prevalence is substantial: precisely when susceptible depletion (included only in the Kendall formulation) matters. Hence, we believe that the differences between the Whittle and Kendall escape pressures arise from susceptible depletion that actually matters for establishment, justifying the use of the Kendall escape pressure. For more general modelling purposes, it might help to consider an establishment probability that (i) accounts for the ongoing susceptible depletion, but also (ii) permits establishment at early appearance times, even if 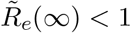 An option could be to calculate the establishment probability using stochastic simulations of the time-inhomogeneous birth-and-death process. Once the mutant strain reaches a threshold prevalence, Gillespie simulations [12] declare success in the establishment process, as in [44]. The crux is specifying this threshold mutant prevalence. For example, [44] uses an arbitrary threshold of 100 infections. Understanding the “First Passage Time (FPT)” [4] of the mutant outbreak—after which stochastic effects are not important—could help find a suitable threshold prevalence. Moreover, the FPT could replace infinity as the limit of the integral in the Kendall formula (9), so that the establishment probability *p*_*K*_(*T* ) only depends on the values of 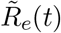 for times *t* preceeding the FPT.

An alternative epidemiological context, however, could mean that strain establishment is really about the longer term persistence of the emerging strain. This scenario occurs, for example, if the epidemic model describes a global pandemic of a pathogen that does not induce lifelong immunity: the survival at low numbers of the mutant strain is important as it measures its potential to generate an other epidemic wave after population immunity has waned. In this scenario, the Kendall formulation correctly captures strain establishment, regardless of the emergence time.

### 4.4 Conclusion

Here we have found an unexpected and new pattern: the overall escape pressure—for the appearance and establishment of escape strains—might be lowest at intermediate vaccination coverages. Whether this “U-shape” pattern appears depends on the strain cross-immunity, which here does not necessarily offer full protection against the mutant. Nevertheless, wildtype infections do shrink the pool of susceptible individuals and may prevent the establishment of the mutant. In general, the escape pressure rate has its maximum shortly before the peak prevalence of the wildtype strain, meaning this is the period of highest risk for immune escape. The implications for monitoring the emergence of new variants during an outbreak is that surveillance efforts (e.g., genomic sequencing of wastewater or sentinel samples) should not be “saved” for the peak of the outbreak (when one might have a priori expected a higher risk of pathogen evolution). Instead, surveillance efforts will be more cost-effective if deployed before the epidemic peak, when any new variants pose greatest threat. Since we account for the depletion of susceptibles that occurs *after* the mutant appears (using a time-inhomogeneous branching process), the results here differ qualitatively from the findings of previous works. In particular, these results show that, statistically, more emerging variants will fail to produce an epidemic wave—despite having an effective reproduction number above one—than would be predicted by classical branching theory.

## Data Availability

The manuscript has no data.

## Acknowledgements

MAG was supported by the Gates Cambridge Trust (grant OPP1144 from the Bill & Melinda Gates Foundation).

## Statements and Declarations

We declare we have no competing interests.

## Appendix: A train interference during infection

The main text assumes that an active infection with the wildtype strain does not protect against the mutant. Therefore, the above results assume that coinfections of hosts with both strains are possible. Coinfections may have substantial consequences for vaccine-induced strain replacement at the population level [23, 34], so here we explore an alternative scenario without coinfections (as in [10, 24, 40]). Excluding coinfections, we find interesting new behaviours (Figure A2) in the establishment probabilities, which further highlight the importance of the Kendall formulation. We found no meaningful differences in the cumulative escape pressures (not shown).

We now assume that wildtype infections temporarily protect hosts against the mutant, due to ecological (e.g., quarantine) [42] or immunological [7] interference. The susceptibility of hosts after recovery from the wildtype is as in the main text. We also maintain the previous assumptions for the relative infectiousness of each host type. Thus, the effective reproduction number of the mutant strain (2) is now

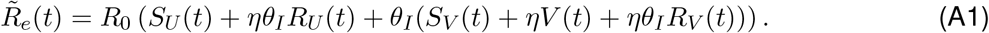

Using 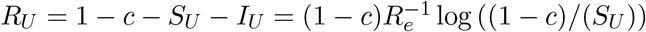 from [18], we get

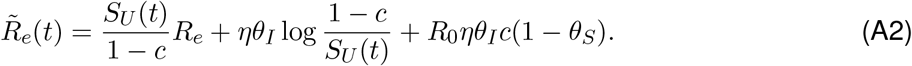

At the start (*t* = 0) and end (*t* ) of the wildtype epidemic, coinfections are impossible even without strain interference—there are no wildtype infections—so at such times 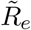 is as before. However, the lack of coinfections at times *t* (0, ) means that 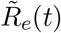 does not necessarily decrease monotonically with time. To understand how 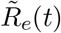 changes with time, we differentiate (A2):

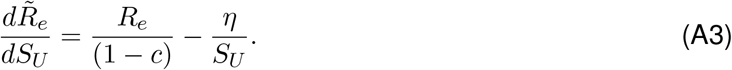

When *S*_*U*_ = 1 − *c*, the derivative (A3) is positive. Since *S*_*U*_ (*t*) is a decreasing function of time, 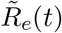 also decreases initially (as *t* increases from zero). The minimum of (A3) is when *S*_*U*_ is smallest,at the end of the epidemic 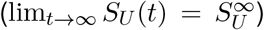 Hence, if 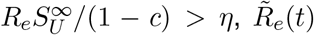 decreases throughout the epidemic (but not necessarily below one), as before. In contrast, if 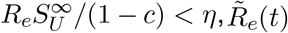 has a local minimum (which may or may not be below one). Hence, for low cross-immunity, the effective reproduction number of the mutant initially decreases (due to wildtype infections, which do not permit coinfection with the mutant) but later increases (because recovered individuals become partially susceptible to the mutant).

Figure A1 shows how the non-monotonicity in 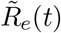 (here due to strain interference) has consequences for the establishment probabilities *p*_*W*_ and *p*_*K*_, relative to the main text (where 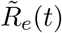 decreases monotonically with time). In the middle (*c* = 50%) and right (*c* = 100%) columns in Figure A1, 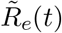 decreases monotonically, eventually dropping below one 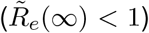 for *c* = 100% but not for *c* = 50%. These plots are virtually indistinguishable from the analogous plots in Figure 1. However, in the left column (*c* = 0%) in Figure A1, 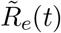 temporarily drops below one but later stabilises above one (as infected hosts recover and rejoin the pool of susceptibles to the mutant). The Whittle establishment probability *p*_*W*_ (*t*) vanishes during the period of 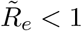 In contrast, the Kendall probability *p*_*K*_(*t*) remains positive. The mutant may sustain enough transmission to persist (in low numbers), “waiting” for establishment once 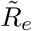 eventually raises above one (because here 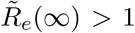 otherwise,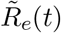 remains permanently below one so the establishment probabilities behave as in the middle columns in Figures 1 and A1). In other words, the mutant may succeed at establishment (*p*_*K*_(*T* ) > 0), even if its effective reproduction number is less than one when it appears 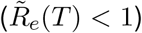 Interestingly, if the mutant appears before the period of 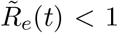 the Kendall probability *p*_*K*_(*T* ) accounts for the temporary drop in 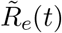, which complicates establishment. Hence, *p*_*K*_(*t*) is lower than *p*_*W*_ (*t*), because *p*_*W*_ does not foresee the upcoming reduction of the pool of susceptibles (as in the other plots and the main text). After the period of 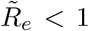 the Kendall probability is higher than the Whittle probability (as proven in Section 2.2), because again *p*_*K*_(*T* ) “sees” that 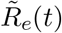 keeps increasing for *t* > *T* . Thus, *P*_*K*_(*T* ) is also greater than *P*_*W*_ (*T* ), but this late in the epidemic both escape pressure rates are close to zero.

**Figure A1:**
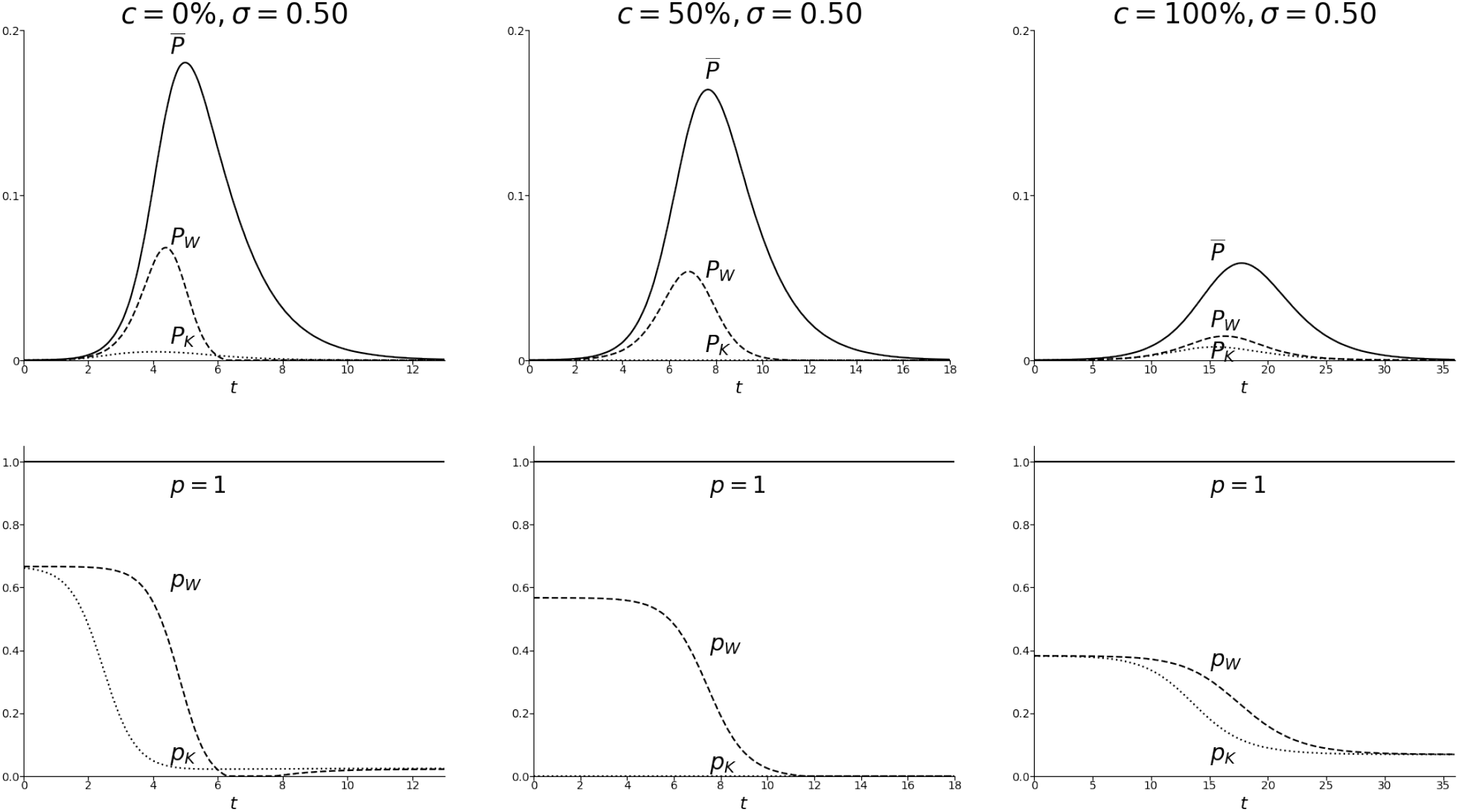
As in Figure 1, but under direct strain interference—active wildtype infections prevent infection with the mutant strain—as per (A1). Top row: escape pressure rates P_⊗_(t) and 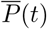. Bottom row: establishment probabilities p_⊗_(t), including p = 1 for comparison. The difference between the columns is the vaccination coverage c, which increases from left (no vaccination) to right (fully vaccinated population). Fixed R_0_ = 3, θ_S_ = 0.8, θ_I_ = 0.6, σ = 0.5, θ_E_ = 7/3 (all values as in Figure 1).

It is possible that *R*_*e*_(*t*) is not monotonic 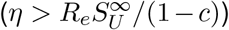 but always remains above one. The local minimum of (A1) occurs—if it exists—when *S*_*U*_ = (1 − *c*)*η/R*_*e*_ and takes the value

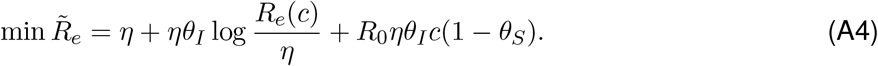

In the left column in Figure A1, min 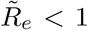 (since *p*_*W*_ = 0 for some intermediate times). However, this minimum (A4) may not always be below one. The expression (A4) is an increasing function of both the vaccination coverage *c* and the immune evasiveness *η* = 1− σ. Therefore, increasing the vaccination coverage or reducing the cross-immunity σ might lead to min 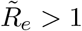 Figure A2 shows two such possibilities (middle and right columns). Since 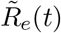 is still non-monotonic, *p*_*W*_ (*t*) also has a temporary decrease, but now remains positive—so establishment is possible—for all appearance times. In this situation, the Kendall probability may still be monotonically decreasing (middle column) or may also experience a temporary drop (right column). The temporary drop in *p*_*K*_ occurs because the strain cross-immunity is low. Low cross-immunity means that the mutant has a relatively high chance of establishment *after* enough hosts recover from the wildtype. However, the mutant strains that appear during the peak in wildtype prevalence are relatively less likely to reach establishment, even though the Kendall probability foresees this upcoming increase in the susceptible pool. Since 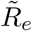 substantially increases after the wildtype peak, the Kendall probability is also non-monotonic. Nevertheless, the temporary drop in *p*_*K*_(*t*) is relatively small. Numerical simulations do not show a qualitative change in the shape of the Kendall escape pressure rate *P*_*K*_(*t*) due to the drop in the establishment probability *p*_*K*_(*t*): *P*_*K*_(*t*) maintains a single local maximum (which now occurs after the peak of *P*_*W*_ (*t*)).

**Figure A2:**
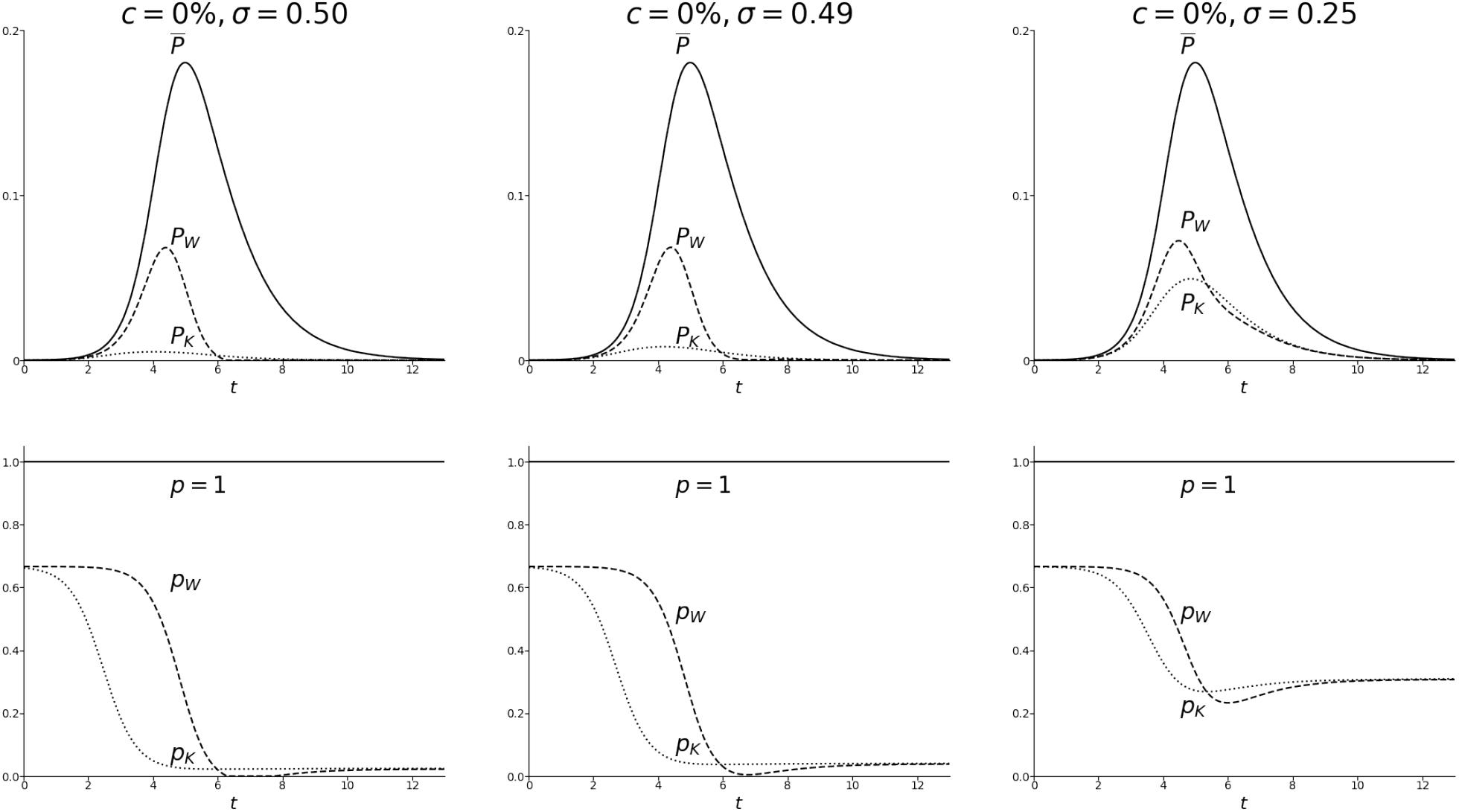
Escape pressure rates P_⊗_(t) and 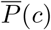 (top row) and establishment probabilities p_⊗_(t) (bottom row, including p = 1 for comparison) with direct strain interference, as in Figure A1. Fixed vaccination coverage c = 0 as in the left column in Figure A1 (repeated here). The cross-immunity σ is lower here in the middle and right columns. Fixed R_0_ = 3, θ_S_ = 0.8, θ_I_ = 0.6, θ_E_ = 7/3 (as in Figure A1) and c = 0.

## References

[1] Arinaminpathy, N., Riley, S., Barclay, W. S., Saad-Roy, C., and Grenfell, B. “Population implications of the deployment of novel universal vaccines against epidemic and pandemic influenza”. Journal of The Royal Society Interface 17.164 (2020), 20190879.

[2] Boven, M. van, Mooi, F. R., Schellekens, J. F., Melker, H. E. de, and Kretzschmar, M. “Pathogen adaptation under imperfect vaccination: implications for pertussis”. Proceedings of the Royal Society B: Biological Sciences 272.1572 (2005), 1617–1624.

[3] Carmona, P. and Gandon, S. “Winter is coming: Pathogen emergence in seasonal environments”. PLOS Computational Biology 16.7 (2020), e1007954.

[4] Curran-Sebastian, J., Pellis, L., Hall, I., and House, T. “Calculation of Epidemic First Passage and Peak Time Probability Distributions”. SIAM/ASA Journal on Uncertainty Quantification 12.2 (2024), 242–261.

[5] Day, T., Galvani, A., Struchiner, C., and Gumel, A. “The evolutionary consequences of vaccination”. Vaccine. The Evolutionary Consequences of Vaccination 26 (2008), C1–C3.

[6] Day, T., Kennedy, D. A., Read, A. F., and Gandon, S. “Pathogen evolution during vaccination campaigns”. PLOS Biology 20.9 (2022), e3001804.

[7] Dietz, K. “Epidemiologic interference of virus populations”. Journal of Mathematical Biology 8.3 (1979), 291–300.

[8] Epstein, S. L. and Price, G. E. “Cross-protective immunity to influenza A viruses”. Expert Review of Vaccines 9.11 (2010), 1325–1342.

[9] Gandon, S. and Day, T. “The evolutionary epidemiology of vaccination”. Journal of The Royal Society Interface 4.16 (2007), 803–817.

[10] Gandon, S., Lambert, A., Voinson, M., Day, T., and Parsons, T. L. “The speed of vaccination rollout and the risk of pathogen adaptation”. medRxiv preprint (2024), 2022.08.01.22278283.

[11] Garcia-Beltran, W. F., Lam, E. C., Denis, K. S., Nitido, A. D., Garcia, Z. H., Hauser, B. M., Feldman, J., Pavlovic, M. N., Gregory, D. J., Poznansky, M. C., Sigal, A., Schmidt, A. G., Iafrate, A. J., Naranbhai, V., and Balazs, A. B. “Multiple SARS-CoV-2 variants escape neutralization by vaccine-induced humoral immunity”. Cell 184.9 (2021), 2372–2383.e9.

[12] Gillespie, D. T. “Stochastic Simulation of Chemical Kinetics”. Annual Review of Physical Chemistry 58 (2007), 35–55.

[13] Gog, J. R. “The impact of evolutionary constraints on influenza dynamics”. Vaccine. Supplement 3 26 (2008), C15– C24.

[14] Gog, J. R., Hill, E. M., Danon, L., and Thompson, R. N. “Vaccine escape in a heterogeneous population: insights for SARS-CoV-2 from a simple model”. Royal Society Open Science 8.7 (2021), 210530.

[15] Gog, J. R., Pellis, L., Wood, J. L. N., McLean, A. R., Arinaminpathy, N., and Lloyd-Smith, J. O. “Seven challenges in modeling pathogen dynamics within-host and across scales”. Epidemics. Challenges in Modelling Infectious Disease Dynamics 10 (2015), 45–48.

[16] Gog, J. R., Rimmelzwaan, G. F., Osterhaus, A. D. M. E., and Grenfell, B. T. “Population dynamics of rapid fixation in cytotoxic T lymphocyte escape mutants of influenza A”. Proceedings of the National Academy of Sciences 100.19 (2003), 11143–11147.

[17] Grenfell, B. T., Pybus, O. G., Gog, J. R., Wood, J. L. N., Daly, J. M., Mumford, J. A., and Holmes, E. C. “Unifying the Epidemiological and Evolutionary Dynamics of Pathogens”. Science 303.5656 (2004), 327–332.

[18] Gutierrez, M. A. and Gog, J. R. “The importance of vaccinated individuals to population-level evolution of pathogens”. Journal of Theoretical Biology 567 (2023), 111493.

[19] Halloran, M. E., Longini Jr., I. M., and Struchiner, C. J. “Design and Interpretation of Vaccine Field Studies”. Epidemiologic Reviews 21.1 (1999), 73–88.

[20] Hartfield, M. and Alizon, S. “Epidemiological Feedbacks Affect Evolutionary Emergence of Pathogens”. The American Naturalist 183.4 (2014).

[21] He, X.-S., Holmes, T. H., Zhang, C., Mahmood, K., Kemble, G. W., Lewis, D. B., Dekker, C. L., Greenberg, H. B., and Arvin, A. M. “Cellular Immune Responses in Children and Adults Receiving Inactivated or Live Attenuated Influenza Vaccines”. Journal of Virology 80.23 (2006), 11756–11766.

[22] Hu, J., Peng, P., Cao, X., Wu, K., Chen, J., Wang, K., Tang, N., and Huang, A.-l. “Increased immune escape of the new SARS-CoV-2 variant of concern Omicron”. Cellular & Molecular Immunology 19.2 (2022), 293–295.

[23] Iannelli, M., Martcheva, M., and Li, X.-Z. “Strain replacement in an epidemic model with super-infection and perfect vaccination”. Mathematical Biosciences 195.1 (2005), 23–46.

[24] Karamitsou, V. “A cross-scale model for the evolution of influenza within a single season”. University of Cambridge. PhD thesis (2021).

[25] Keeling, M. J. and Rohani, P. Modeling Infectious Diseases in Humans and Animals. Princenton University Press, 2007.

[26] Kendall, D. G. “On the Generalized “Birth-and-Death” Process”. The Annals of Mathematical Statistics 19.1 (1948), 1–15.

[27] Kissler, S. M., Hay, J. A., Fauver, J. R., Mack, C., Tai, C. G., Anderson, D. J., Ho, D. D., Grubaugh, N. D., and Grad, Y. H. “Viral kinetics of sequential SARS-CoV-2 infections”. Nature Communications 14.1 (2023), 6206.

[28] Kissler, S. M. et al. “Viral Dynamics of SARS-CoV-2 Variants in Vaccinated and Unvaccinated Persons”. New England Journal of Medicine 385.26 (2021), 2489–2491.

[29] Kucharski, A. and Gog, J. R. “Influenza emergence in the face of evolutionary constraints”. Proceedings of the Royal Society B: Biological Sciences 279.1729 (2011), 645–652.

[30] Lehtonen, J. “The Lambert W function in ecological and evolutionary models”. Methods in Ecology and Evolution 7.9 (2016), 1110–1118.

[31] Liu, J., Yu, Y., Jian, F., Yang, S., Song, W., Wang, P., Yu, L., Shao, F., and Cao, Y. “Enhanced immune evasion of SARS-CoV-2 variants KP.3.1.1 and XEC through N-terminal domain mutations”. The Lancet Infectious Diseases 25.1 (2025), e6–e7.

[32] Lloyd-Smith, J. O., Schreiber, S. J., Kopp, P. E., and Getz, W. M. “Superspreading and the effect of individual variation on disease emergence”. Nature 438.7066 (2005), 355–359.

[33] Lobinska, G., Pauzner, A., Traulsen, A., Pilpel, Y., and Nowak, M. A. “Evolution of resistance to COVID-19 vaccination with dynamic social distancing”. Nature Human Behaviour 6.2 (2022), 193–206.

[34] Martcheva, M., Bolker, B. M., and Holt, R. D. “Vaccine-induced pathogen strain replacement: what are the mechanisms?” Journal of The Royal Society Interface 5.18 (2007), 3–13.

[35] Moss, P. “The T cell immune response against SARS-CoV-2”. Nature Immunology 23.2 (2022), 186–193.

[36] Nielsen, B. F., Saad-Roy, C. M., Metcalf, C. J. E., Viboud, C., and Grenfell, B. T. “Eco-evolutionary dynamics of pathogen immune-escape: deriving a population-level phylodynamic curve”. Journal of The Royal Society Interface 22.225 (2025), 20240675.

[37] Park, A. W., Daly, J. M., Lewis, N. S., Smith, D. J., Wood, J. L. N., and Grenfell, B. T. “Quantifying the Impact of Immune Escape on Transmission Dynamics of Influenza”. Science 326.5953 (2009), 726–728.

[38] Rahman, S., Rahman, M. M., Miah, M., Begum, M. N., Sarmin, M., Mahfuz, M., Hossain, M. E., Rahman, M. Z., Chisti, M. J., Ahmed, T., Arifeen, S. E., and Rahman, M. “COVID-19 reinfections among naturally infected and vaccinated individuals”. Scientific Reports 12.1 (2022), 1438.

[39] Rella, S. A., Kulikova, Y. A., Minnegalieva, A. R., and Kondrashov, F. A. “Complex vaccination strategies prevent the emergence of vaccine resistance”. Evolution 78.10 (2024), 1722–1738.

[40] Rella, S. A., Kulikova, Y. A., Dermitzakis, E. T., and Kondrashov, F. A. “Rates of SARS-CoV-2 transmission and vaccination impact the fate of vaccine-resistant strains”. Scientific Reports 11.1 (2021), 15729.

[41] Restif, O. and Grenfell, B. T. “Vaccination and the dynamics of immune evasion”. Journal of The Royal Society Interface 4.12 (2006), 143–153.

[42] Rohani, P., Earn, D. J., Finkenstädt, B., and Grenfell, B. T. “Population dynamic interference among childhood diseases”. Proceedings of the Royal Society of London. Series B: Biological Sciences 265.1410 (1998), 2033–2041.

[43] Saad-Roy, C. M., Morris, S. E., Metcalf, C. J. E., Mina, M. J., Baker, R. E., Farrar, J., Holmes, E. C., Pybus, O. G., Graham, A. L., Levin, S. A., Grenfell, B. T., and Wagner, C. E. “Epidemiological and evolutionary considerations of SARS-CoV-2 vaccine dosing regimes”. Science 372.6540 (2021), 363–370.

[44] Sachak-Patwa, R., Byrne, H. M., Dyson, L., and Thompson, R. N. “The risk of SARS-CoV-2 outbreaks in low prevalence settings following the removal of travel restrictions”. Communications Medicine 1.1 (2021), 1–9.

[45] Subramanian, R., Graham, A. L., Grenfell, B. T., and Arinaminpathy, N. “Universal or Specific? A Modeling-Based Comparison of Broad-Spectrum Influenza Vaccines against Conventional, Strain-Matched Vaccines”. PLOS Computational Biology 12.12 (2016), e1005204.

[46] Tarke, A. et al. “Impact of SARS-CoV-2 variants on the total CD4+ and CD8+ T cell reactivity in infected or vaccinated individuals”. Cell Reports Medicine 2.7 (2021).

[47] Thompson, R. N., Hill, E. M., and Gog, J. R. “SARS-CoV-2 incidence and vaccine escape”. The Lancet Infectious Diseases 21.7 (2021), 913–914.

[48] Thompson, R. N., Southall, E., Daon, Y., Lovell-Read, F. A., Iwami, S., Thompson, C. P., and Obolski, U. “The impact of cross-reactive immunity on the emergence of SARS-CoV-2 variants”. Frontiers in Immunology 13 (2023).

[49] Whittle, P. “The outcome of a stochastic epidemic—a note on Bailey’s paper”. Biometrika 42.1 (1955), 116–122.

[50] Zarnitsyna, V. I., Bulusheva, I., Handel, A., Longini, I. M., Halloran, M. E., and Antia, R. “Intermediate levels of vaccination coverage may minimize seasonal influenza outbreaks”. PLOS One 13.6 (2018), e0199674.

[51] Zhang, X.-S., Pebody, R., Angelis, D. D., White, P. J., Charlett, A., and McCauley, J. W. “The Possible Impact of Vaccination for Seasonal Influenza on Emergence of Pandemic Influenza via Reassortment”. PLOS One 9.12 (2014), e114637.

[52] Zhang, X., Lobinska, G., Feldman, M., Dekel, E., Nowak, M. A., Pilpel, Y., Pauzner, Y., Barzel, B., and Pauzner, A. “A spatial vaccination strategy to reduce the risk of vaccine-resistant variants”. PLOS Computational Biology 18.8 (2022), e1010391.

[53] Zinder, D., Bedford, T., Gupta, S., and Pascual, M. “The Roles of Competition and Mutation in Shaping Antigenic and Genetic Diversity in Influenza”. PLOS Pathogens 9.1 (2013), e1003104.

